# Emerging Barriers to Accessing Cataract Surgery in a Low- and Middle-Income Country: A Hospital-Based Cross-Sectional Study from Bangladesh

**DOI:** 10.1101/2025.11.26.25341107

**Authors:** Pradipta N. Chowdhury, Altaf U. Khan, Yasmin Akter, Sk. Mohammad Murad, Anindita Chowdhury, Minhaz Uddin, Priya Mitra, Prakash K. Chowdhury

## Abstract

**Purpose:** To assess the key barriers that limit access to cataract surgery services in the lower-middle-income country, Bangladesh, with a high cataract burden.

**Methods:** A cross-sectional preoperative survey was conducted among 595 cataract patients attending hospital outpatient services (n = 222) and community outreach camps (n = 373) in Lions Charitable Eye Hospital from May to August 2025. Data were collected using systematic sampling through an interviewer-administered questionnaire. Socio-demographic characteristics, awareness of cataracts and treatment, and barriers to surgery were assessed. Regression analyses were performed to evaluate the associations between patient characteristics and the reported barriers.

**Results:** The study included 595 cataract patients (mean age 62 years; 50% women). Most of them presented with bilateral cataracts (74%). Camp patients were more often rural and less educated (p < 0.001). Although most patients reporting barriers had heard of cataract (72.4**%**), fewer knew about treatment options (38.2%), with knowledge strongly linked to education (p < 0.01). The leading barriers were affordability (430, 72.3%), insufficient time due to work or family responsibilities (165, 27.7%), worry about surgical outcomes (161, 27.1%), fear of the procedure (154, 25.9%), transportation (138, 23.2%), and lack of escort (131, 22%). Women frequently cited fear and lack of escort (p < 0.001) as barriers. Lower education and income were associated with more barriers, while prior surgery was found to reduce them (Adj. R² = 0.138). Cataract-related visual impairment significantly limited daily functioning, particularly household work (47.9%) and mobility (45.4%).

**Conclusion:** Financial, socio-cultural, and accessibility barriers limit uptake of cataract surgery in Bangladesh. The existing disparities in access to cataract surgery can be addressed through health promotion, medical staff training, and enhanced patient counseling.

## 1. Introduction

Globally, over 2.2 billion individual live with near or distant visual impairment, 1 billion of them being preventable [1]. South Asia has a higher cataract-associated blindness burden compared to the global average [2]. The highly populated South Asian country, Bangladesh, has over 6 million visually impaired and 0.75 million blind individuals [3].

Cataract causes distance vision impairment or blindness in an estimated 94 million people [1]. A cataract occurs when the eye’s natural lens becomes cloudy due to the breakdown of proteins, causing blurred or hazy vision [4]. Cataract-induced visual impairment unequally affects women, members from financially disadvantaged households, and populations residing in low and middle-income countries (LMICs) [5]. Technological advances have made cataract surgery more efficacious, but access remains inadequate in rural areas of developing nations with a high vision loss prevalence [6].

Accessibility is a critical social determinant of visual health, comprising equality, physical access, affordability, and access to information [1]. Health inequities arise when social, economic, and structural factors lead to uneven distribution of resources, causing disparities among medically underserved groups [7, 8]. Ensuring the availability of services only partly addresses the global challenge of visual impairment. It is equally critical to tackle social, cultural, and informational barriers that prevent the uptake of cataract surgery [2, 9].

The commonly cited barriers to eye care in developing countries include cost, fear of surgery, and transportation [10–13]. A cross-sectional study in New Zealand found that longer driving distances were a geographical barrier to accessing ophthalmic care for the Maori ethnicity [14]. Multiple studies in the US showcase that low income, lower education, female gender are associated with increased cataract prevalence [8]. However, barriers observed in developed countries may not be comparable to those in Bangladesh, which has unique socio-cultural, demographic, and economic factors and limited eye care resources for its large population. Limited knowledge, low awareness, and financial constraints were identified as significant barriers to accessing appropriate healthcare in a cross-sectional mixed-methods study among children in the Shatkhira district of Bangladesh [15]. Additionally, the Rohingya refugee community in Bangladesh reported ‘no perceived need’ and ‘expenses’ as the most common barriers to cataract surgery for untreated bilateral cataract [16]. These barrier studies, however, are population-specific, limiting generalization to all subgroups of the population.

Although cataract prevalence is higher among females in Bangladesh, they are less likely than men to undergo cataract surgery. This gap is shaped by patriarchal norms that prioritize men’s health needs and restrict women’s autonomy in making healthcare decisions. Factors such as financial dependence, limited physical access, and cultural stigma further amplify these disadvantages [2, 17]. Additionally, pre-operative fear, driven by local rumors of poor surgical outcomes and pain during surgery, acts as a key barrier to undergoing surgery. Low literacy rates may facilitate the spread of such word-of-mouth misinformation, reinforcing fear within the community [18].

Evidence to inform strategies for enhancing cataract surgery uptake in low and middle-income countries remains scarce. The primary goal of the current study is to identify emerging health system and community-level barriers in a lower-middle-income country, such as Bangladesh, across different population groups of cataract patients.

## 2. Methods

### 2.1 Study Design and Context

The Lions Charitable Eye Institute and Hospital organizes hospital-based outpatient services and community-based “outreach camps” in remote areas of Chattogram, employing their own staff and equipment.

Patients attending the outpatient facility pay a registration fee of 200 BDT (approximately 1.64 USD). Patients diagnosed and advised for cataract surgery are scheduled for a surgery date and asked to return. Those who can afford the procedure pay a fee, while others receive subsidized support from the hospital and its partner organizations. The hospital organizes 10-15 health outreach camps every month in rural and marginalized communities, collaborating with local organizations and community groups. At the outreach camp, patient registration is free, and any subsequent cataract surgery is also provided at no cost. Later, the hospital van carries the patients diagnosed with cataracts to the hospital. After arriving at the hospital, an optometrist performs the initial vision check-up and diagnoses a spectacle based on the required refractive correction. If the spectacles do not improve vision, the patients are recommended to an ophthalmologist. The ophthalmologist advises cataract patients to undergo surgery after evaluating Random Blood Sugar (RBS), Electrocardiogram (ECG), Skin Prick Test (SPT), B-scan ultrasonography, and blood pressure. Patients facing other visual comorbidities, such as diabetic retinopathy, age-related macular degeneration, and glaucoma are recommended to schedule surgery later.

The cross-sectional study was conducted through a preoperative survey of patients undergoing cataract surgery over a period of 4 months from May 2025 to August 2025.

### 2.2 Participants and Data Collection

Hospital facilities and outreach camps were preselected, and patients were systematically sampled from those attending either setting on the designated data collection days.

The sample size was determined using a single-proportion formula to estimate the prevalence of barriers to cataract surgery. A prevalence of 50% was assumed to maximize the required sample size, with a 95% confidence level and 5% margin of error. The minimum required sample size was calculated as 385 participants. The higher-than-expected patient turnout during data collection ultimately led to exceeding this minimum at all study sites. At each hospital facility and outreach camp, a sampling interval (X), ranging from 1 to 10, was calculated by dividing the average daily patient volume by the target sample size. Every Xth patient was then invited to participate. Data were collected over four months, and because attendance was higher than expected, the minimum required sample size was exceeded at all sites.

The data were collected using an interviewer-administered Google form and a hard-copy questionnaire. The questionnaire was translated into Bangla. One research coordinator and two medical officers carried out the data collection throughout the study period. The data collectors were trained beforehand regarding the administration of the questionnaire, interview techniques, and use of the Google Form application. The data collectors performed a pilot test post-training among the hospital patients. Responses were collected using a touchscreen, keyless, ultra-portable application on the mobile phones.

The study included patients with operable cataract and scheduled for surgery. Participants with high blood pressure, high diabetes, abnormal retinal pathology, optic nerve, and macular morbidities were excluded from the study. Patients too unwell to speak were also excluded from the study.

### 2.3 Ethics Statement

The study adhered to the principles outlined in the Declaration of Helsinki. The Lions Charitable Eye Institute Ethics Review Board granted ethical approval. Each study was conducted in line with the fundamental principles of health research, centered on the commitment to avoid harm. Participants were thoroughly informed about the study prior to the interview and were assured of confidentiality. Verbal and written consent were sought and recorded. While adults who had difficulty communicating or understanding were being interviewed, verbal consent was requested from an accompanying family member or caregiver. If participants refused to give consent, the interview was not pursued further.

## 3. Results

### 3.1 Socio-demographics and Clinical Characteristics

The cross-sectional study included a total of 595 patients, comprising 373 camp patients and 222 outpatients. The study participants’ ages ranged from 25 to 90 years old. The mean age was 62 years (SD 10), and the gender distribution was balanced, with women representing just over half of the sample (50.3%).

**Table 1** presents the socio-demographic characteristics of the study participants, highlighting the apparent differences between camp and OPD patients.

**Table 1.** Baseline Socio-Demographic and Clinical Characteristics of the Study Population.

| *Characteristic* | *Total Cohort (N=595)* | *Camp Patients (n=373)* | *OPD Patients (n=222)* | *p-value* |
| --- | --- | --- | --- | --- |
| **Gender** |  |  |  | 0.304 |
| Female | 299 (50.3%) | 194 (52.0%) | 105 (47.3%) |  |
| Male | 296 (49.7%) | 179 (48.0%) | 117 (52.7%) |  |
| **Location** |  |  |  | <0.001 |
| Rural | 354 (59.5%) | 284 (76.1%) | 70 (31.5%) |  |
| Urban | 138 (23.2%) | 41 (11.0%) | 97 (43.7%) |  |
| Semi-urban | 94 (15.8%) | 39 (10.5%) | 55 (24.8%) |  |
| Ethnic minority | 9 (1.5%) | 9 (2.4%) | 0 (0.0%) |  |
| **Education** |  |  |  | <0.001 |
| Class 5 or less | 214 (36.0%) | 135 (36.2%) | 79 (35.6%) |  |
| Never been to school | 176 (29.6%) | 153 (41.0%) | 23 (10.4%) |  |
| HSC or General Diploma | 58 (9.7%) | 16 (4.3%) | 42 (18.9%) |  |
| 4-year college graduate | 10 (1.7%) | 3 (0.8%) | 7 (3.2%) |  |
| More than 4-year degree | 5 (0.8%) | 0 (0.0%) | 5 (2.3%) |  |
| **Occupation** |  |  |  | 0.012 |
| Housewife | 266 (44.7%) | 177 (47.5%) | 89 (40.1%) |  |
| Retired | 119 (20.0%) | 75 (20.1%) | 44 (19.8%) |  |
| Employed for wages | 87 (14.6%) | 44 (11.8%) | 43 (19.4%) |  |
| Self-employed | 68 (11.4%) | 40 (10.7%) | 28 (12.6%) |  |
| Other/Unemployed | 55 (9.3%) | 37 (9.9%) | 18 (8.1%) |  |
| **Spectacle use** |  |  |  | 0.152 |
| Yes | 174 (29.2%) | 98 (26.3%) | 76 (34.2%) |  |
| No | 421 (70.8%) | 275 (73.7%) | 146 (65.8%) |  |
| **Last eye exam** |  |  |  | 0.042 |
| Within 1 year | 157 (26.4%) | 83 (22.3%) | 74 (33.3%) |  |
| More than 1 year | 438 (73.6%) | 290 (77.7%) | 148 (66.7%) |  |
| **Visual impairment** |  |  |  | <0.001 |
| Unilateral | 216 (36.3%) | 130 (34.9%) | 86 (38.7%) |  |
| Bilateral | 379 (63.7%) | 243 (65.1%) | 136 (61.3%) |  |
| **Prior surgery status** |  |  |  | 0.021 |
| Yes | 144 (24.2%) | 76 (20.4%) | 68 (30.6%) |  |
| No | 451 (75.8%) | 297 (79.6%) | 154 (69.4%) |  |
| **Heard about cataract** |  |  |  | 0.005 |
| Yes | 515 (86.6%) | 312 (83.6%) | 203 (91.4%) |  |
| No | 80 (13.4%) | 61 (16.4%) | 19 (8.6%) |  |
| **Knowledge of treatment options** |  |  |  | 0.032 |
| Yes | 462 (77.6%) | 278 (74.5%) | 184 (82.9%) |  |
| No | 133 (22.4%) | 95 (25.5%) | 38 (17.1%) |  |
| **Income group (USD)** |  |  |  | 0.047 |
| Low (<\$100) | 312 (52.4%) | 217 (58.2%) | 95 (42.8%) | |
| Middle (\$100–300) | 243 (40.8%) | 135 (36.2%) | 108 (48.6%) | |
| High (>\$300) | 40 (6.7%) | 21 (5.6%) | 19 (8.6%) | |
| **Family Members (Mean ± SD)** | 5.2 (2.1) | 5.5 (2.2) | 4.7 (2.0) | <0.001 |
| **Age (Mean ± SD)** | 61.3 (10.2) | 62.8 (9.8) | 59.0 (10.5) | <0.001 |

**Table 2.**
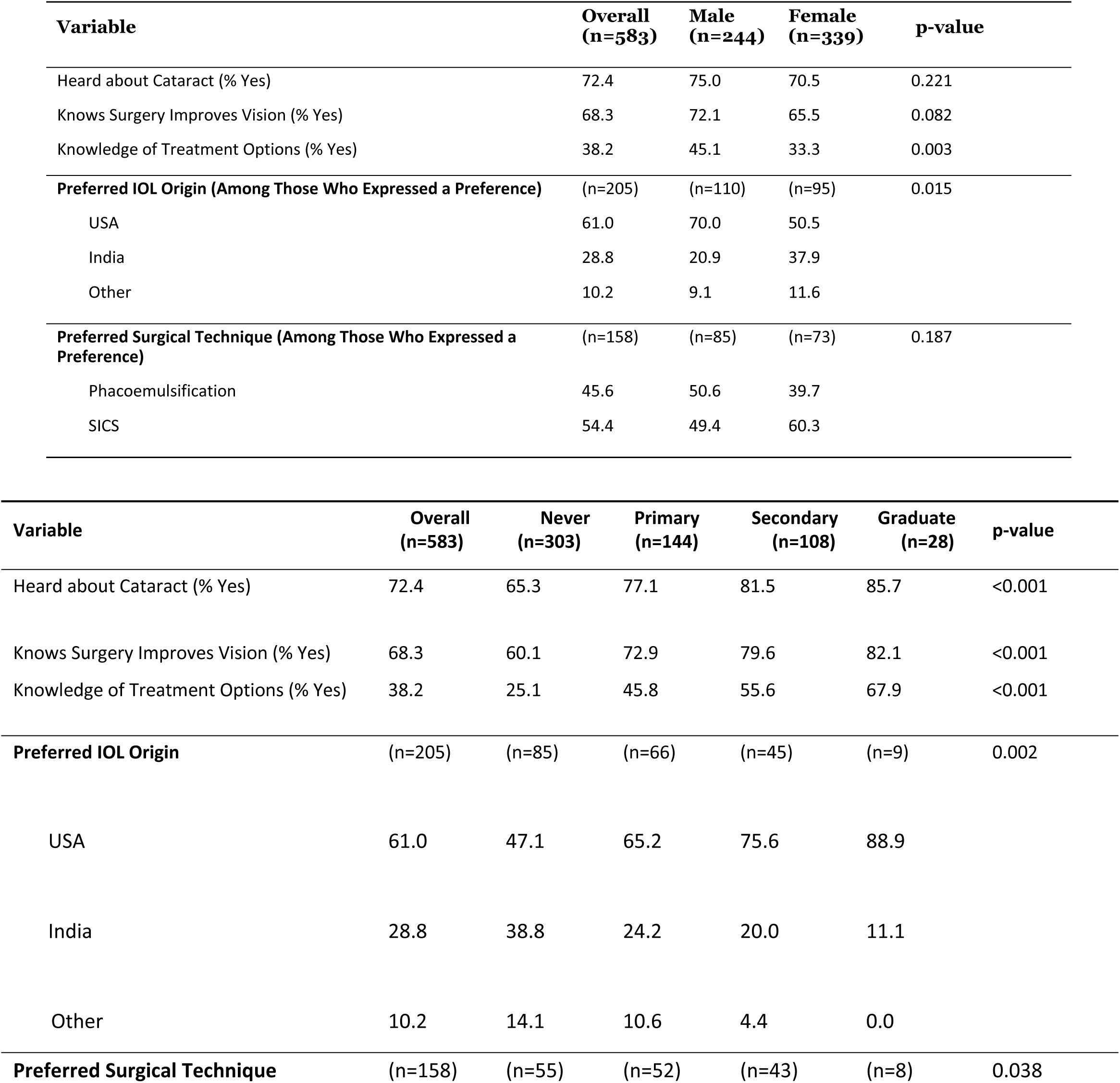
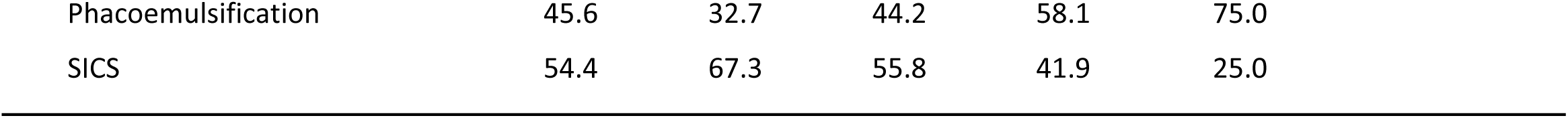
Knowledge and Preferences Regarding Cataract Treatment among Patients Reporting Barriers (N=583)

| Variable | Overall<br>(n=583) | Male<br>(n=244) | Female<br>(n=339) | p-value |
| --- | --- | --- | --- | --- |
| Heard about Cataract (% Yes) | 72.4 | 75.0 | 70.5 | 0.221 |
| Knows Surgery Improves Vision (% Yes) | 68.3 | 72.1 | 65.5 | 0.082 |
| Knowledge of Treatment Options (% Yes) | 38.2 | 45.1 | 33.3 | 0.003 |
| **Preferred IOL Origin (Among Those Who Expressed a Preference)** | (n=205) | (n=110) | (n=95) | 0.015 |
| USA | 61.0 | 70.0 | 50.5 |  |
| India | 28.8 | 20.9 | 37.9 |  |
| Other | 10.2 | 9.1 | 11.6 |  |
| **Preferred Surgical Technique (Among Those Who Expressed a Preference)** | (n=158) | (n=85) | (n=73) | 0.187 |
| Phacoemulsification | 45.6 | 50.6 | 39.7 |  |
| SICS | 54.4 | 49.4 | 60.3 |  |

| Variable | Overall<br>(n=583) | Never<br>(n=303) | Primary<br>(n=144) | Secondary<br>(n=108) | Graduate<br>(n=28) | p-value |
| --- | --- | --- | --- | --- | --- | --- |
| Heard about Cataract (% Yes) | 72.4 | 65.3 | 77.1 | 81.5 | 85.7 | <0.001 |
| Knows Surgery Improves Vision (% Yes) | 68.3 | 60.1 | 72.9 | 79.6 | 82.1 | <0.001 |
| Knowledge of Treatment Options (% Yes) | 38.2 | 25.1 | 45.8 | 55.6 | 67.9 | <0.001 |
| **Preferred IOL Origin** | (n=205) | (n=85) | (n=66) | (n=45) | (n=9) | 0.002 |
| USA | 61.0 | 47.1 | 65.2 | 75.6 | 88.9 |  |
| India | 28.8 | 38.8 | 24.2 | 20.0 | 11.1 |  |
| Other | 10.2 | 14.1 | 10.6 | 4.4 | 0.0 |  |
| **Preferred Surgical Technique** | (n=158) | (n=55) | (n=52) | (n=43) | (n=8) | 0.038 |

|  |  |  |  |  |  |
| --- | --- | --- | --- | --- | --- |
| Phacoemulsification | 45.6 | 32.7 | 44.2 | 58.1 | 75.0 |
| SICS | 54.4 | 67.3 | 55.8 | 41.9 | 25.0 |

In terms of clinical characteristics, bilateral cataract was the most frequent presentation, affecting nearly three-quarters of participants (74%). This finding was more common among OPD patients (80%) than those attending camps (70%) (p = 0.039). 59.9% of females (n = 179) and 57.8% of males (n = 171) presented with bilateral cataracts, not showcasing any statistical significance (Chi-square, p = 0.075). A minority (19%) reported a history of prior cataract surgery, with no significant difference between groups. 91% of the participants who underwent cataract surgery reported knowing someone who had a positive surgical outcome.

### 3.2 Prevalent Knowledge Regarding Cataract Treatment

Patients’ knowledge and awareness of cataracts and available treatment options were assessed through three questions: (1) “Have you heard of cataracts?” (2) “Do you know about the available treatment options?” and (3) “Do you believe surgery can improve vision?” Among the participants who reported barriers, 72.4**%** had heard of cataracts, while 38.2**%** were aware of the available treatment options for cataracts. Additionally, 68.3% perceived that surgery could restore vision. Males had significantly higher knowledge of treatment option than females (45.1% vs. 33.3%, p ≈ 0.003), though no significant gender differences were found in terms of general awareness of cataracts (p = .082) or the belief that surgery improves vision (p ≈ 0.221).

Among the patients who answered IOL factor questions, 96.8% believed the country of manufacture matters more than IOL type/technology (3.2%). Among those stating a preference, 82.2% chose “Made in USA,” 14.5% “Made in India,” and 3.3% “Trifocal.” Among the respondents choosing a technique, 50.9% chose Phacoemulsification, while 49.1% selected SICS. Education had a significant impact on awareness across all items (p < 0.01). 59.8% (n = 356) of the participants undergoing cataract surgery reported knowing someone with a positive surgical experience.

### 3.3 Patient-perceived barriers

Each patient reported multiple barriers to seeing an eye doctor. The fig 1 showcases most commonly claimed barriers, which include medical costs (71.4%), transportation (27.7%), getting time off work (21.3%), getting an appointment (16.6%), and lack of available services (16%), followed by negligence (9.2%) and difficulty navigating the health system (7.1%).

**Fig 1.**
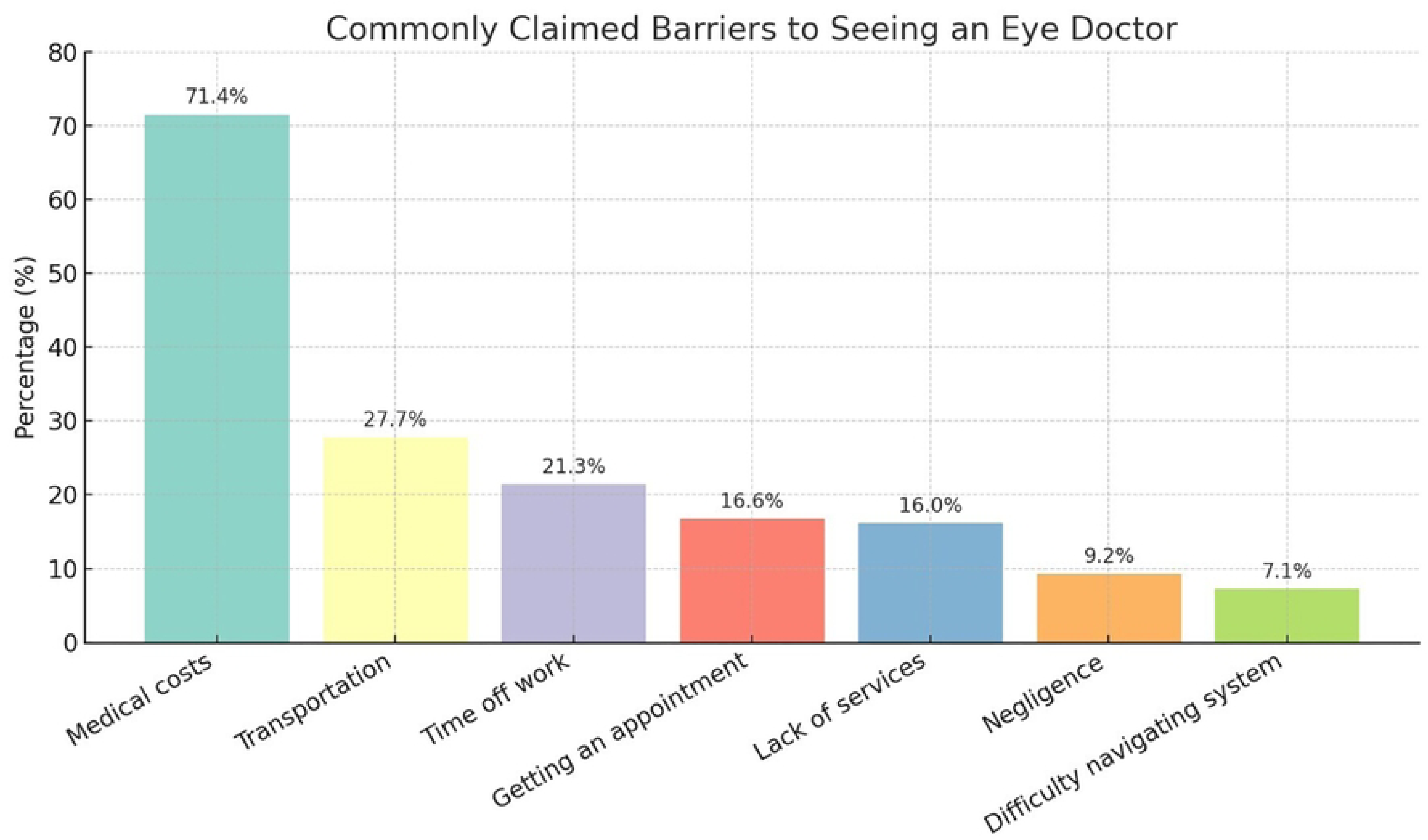
Most Frequently Reported Barriers to Seeing an Eye Doctor. The bar chart shows the percentage of patients reporting each barrier to seeing an eye doctor.

**Fig 2.**
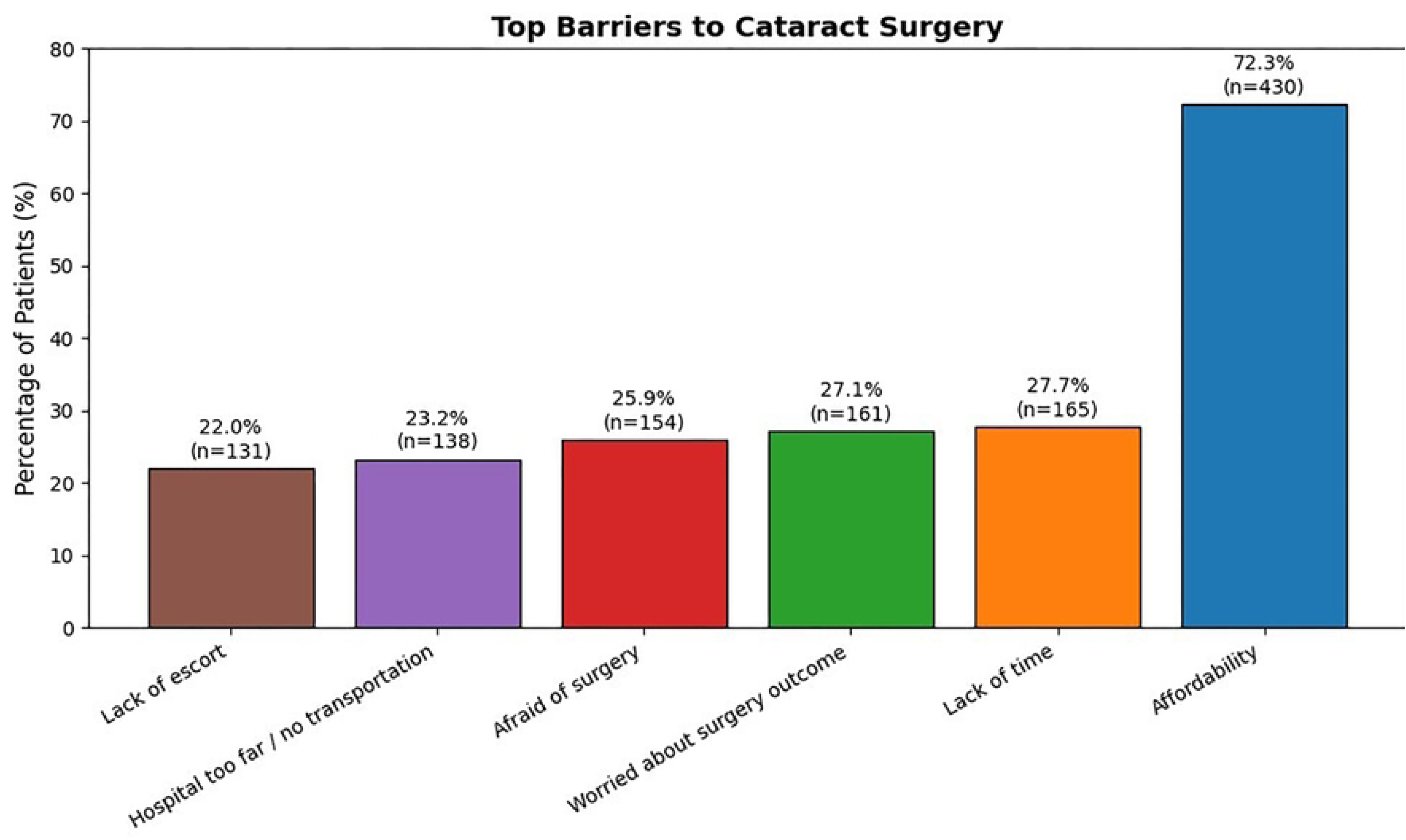
The Top Emerging Barriers to Cataract Surgery. The bar chart shows the percentage of patients reporting each barrier.The most prevalent barriers in our study include affordability (430,72.3%), time constraints due to work or family responsibilities (165, 27.7%), worry about surgical outcomes (161, 27.1%), fear of surgery (154, 25.9%), transportation (138, 23.2%), and lack of escort (131, 22%). The numbers in parentheses indicate the number of patients reporting each barrier.

Many patients shared more than one barrier to accessing cataract surgery. All the reported barriers were considered for percentage calculation to achieve a comprehensive overview of the most frequently reported ones.

The analysis of factors associated with key barriers to cataract surgery revealed several significant associations, as detailed in Table 3. A strong gradient was observed based on socioeconomic status; individuals with lower income levels were significantly more likely to report an inability to afford surgery compared to their higher-income counterparts (79.1% vs. 29.2%, p<0.001). Similarly, cost was a more frequently cited barrier among those with no formal education than among graduates (75.0% vs. 53.3%, p = 0.022).

**Table 3.** Factors Associated with Key Barriers to Cataract Surgery.

| Factor | Category | Barrier |  |  |  |  |  |
| --- | --- | --- | --- | --- | --- | --- | --- |
|  |  | Could not afford | No time | Worried about outcome | Afraid of surgery | No transport | No escort |
| Gender | Male | 220 (73.6) | 91 (30.7) | 40 (16.2) | 80 (27) | 72 (24.3) | 47 (15.9) |
|  | Female | 210 (70.9) | 74 (24.7) | 121 (34.8) | 81 (27.1) | 66 (22.1) | 84 (28.1) |
|  | p-value | 0.532 | 0.123 | <0.001 | 1 | 0.58 | <0.001 |
| Education | Never school | 186 (75.0) | 63 (25.4) | 58 (23.4) | 84 (34.0) | 83 (33.6) | 59 (23.8) |
|  | Primary | 109 (72.9) | 47 (31.4) | 47 (31.4) | 30 (20.1) | 14 (9.4) | 32 (21.4) |
|  | Secondary | 119 (70.6) | 43 (25.5) | 53 (31.4) | 34 (20.1) | 34 (20.1) | 30 (17.8) |
|  | Graduate | 16 (53.3) | 12 (40.0) | 3 (10.0) | 6 (20.0) | 5 (16.7) | 10 (33.3) |
|  | p-value | 0.022 | 0.023 | 0.014 | 0.004 | <0.001 | 0.327 |
| Prior surgery | Yes | 79 (66.4) | 30 (25.2) | 18 (15.1) | 16 (13.4) | 8 (6.7) | 9 (7.6) |
|  | No | 351 (73.9) | 135 (28.4) | 143 (30.1) | 138 (29.0) | 130 (27.4) | 122 (25.6) |
|  | p-value | 0.084 | 0.592 | 0.001 | <0.001 | <0.001 | <0.001 |
| Location | Rural | 321 (71.3) | 100 (22.2) | 121 (26.9) | 125 (27.8) | 121 (27.0) | 100 (22.2) |
|  | Urban | 67 (80.7) | 42 (50.6) | 20 (24.1) | 6 (7.2) | 2 (2.4) | 8 (9.6) |
|  | Semi-urban | 26 (72.2) | 14 (38.9) | 15 (41.7) | 9 (25.0) | 3 (8.3) | 7 (19.4) |
|  | Ethnic | 16 (66.7) | 9 (37.5) | 5 (20.8) | 14 (58.3) | 12 (50.0) | 16 (66.7) |
|  | p-value | 0.001 | <0.001 | <0.001 | 0.184 | <0.001 | <0.001 |
| Income | Low | 315 (79.1) | 97 (24.4) | 121 (30.4) | 128 (32.2) | 116 (29.1) | 98 (24.6) |
|  | Medium | 108 (62.4) | 57 (32.9) | 38 (22.0) | 38 (22.0) | 26 (15.0) | 30 (17.3) |
|  | High | 7 (29.2) | 11 (45.8) | 2 (8.3) | 2 (8.3) | 0 (0.0) | 3 (12.5) |
|  | p-value | <0.001 | 0.014 | 0.012 | <0.001 | <0.001 | 0.041 |
| Age | ≤60 | 194 (70.3) | 101 (36.6) | 62 (22.5) | 57 (20.7) | 58 (21.0) | 50 (18.1) |
|  | >60 | 236 (74.0) | 64 (20.1) | 99 (31.0) | 99 (31.0) | 80 (25.1) | 81 (25.4) |
|  | p-value | 0.316 | <0.001 | 0.019 | 0.007 | 0.241 | 0.049 |

\* Values are n (%). Percentages represent the proportion within each category. P-values from Chi-square or Fisher's exact test.

Notable disparities were identified by gender. Female participants were significantly more likely to express concern about the surgical outcome (34.8% vs. 16.2%, p < 0.001) and to report a lack of an escort (28.1% vs. 15.9%, p < 0.001).

A history of prior surgery is highly associated with a lower prevalence of nearly all barriers. Participants with no prior surgical experience were significantly more likely to report being afraid of surgery (29.0% vs. 13.4%, p<0.001), having no transport (27.4% vs. 6.7%, p<0.001), and having no escort (25.6% vs. 7.6%, p<0.001).

Geographic location also played a prominent role. Urban residents most frequently cited cost (80.7%) and lack of time (50.6%, p<0.001 for overall location comparison), whereas individuals from ethnic minority communities reported the highest rates of being afraid of surgery (58.3%) and faced the most significant logistical challenges, with half citing no transport (50.0%) and two-thirds having no escort (66.7%, p<0.001 for both transport and escort across locations).

Finally, younger participants (≤60 years) were significantly more likely to cite lack of time as a barrier (36.6% vs. 20.1%, p<0.001). In contrast, older participants (>60 years) were more likely to report being worried about the outcome (31.0% vs. 22.5%, p = 0.019), fearful of surgery (31.0% vs. 20.7%, p = 0.007), and lacking an escort (25.4% vs. 18.1%, p = 0.049). Findings indicate that 83.9% of patients had someone to assist with care or medications, 7.4% did not have a caregiver, and 8.2% reported not requiring any assistance.

Fig 3 showcases the associations between patient characteristics and the number of reported barriers to accessing cataract surgery through a linear regression analysis. The model indicated that **e**ducation level and income were significant predictors of the number of barriers. Patients with primary (β = 0.60, p < 0.001) and secondary education (β = 0.65, p < 0.001) reported higher numbers of barriers. Conversely, higher income groups reported fewer barriers: lower-middle (β = -0.23, p = 0.041), upper-middle (β = -0.24, p = 0.033), and high income (β = -0.65, p < 0.001). Prior cataract surgery was associated with a slight reduction in reported barriers (β = -0.19, p = 0.057). Other variables, including age, gender, location, family size, and higher education levels, were not significantly associated with the number of reported barriers. The overall model explained approximately 16% of the variance in barrier counts (Adjusted R² = 0.138).

**Fig 3.**
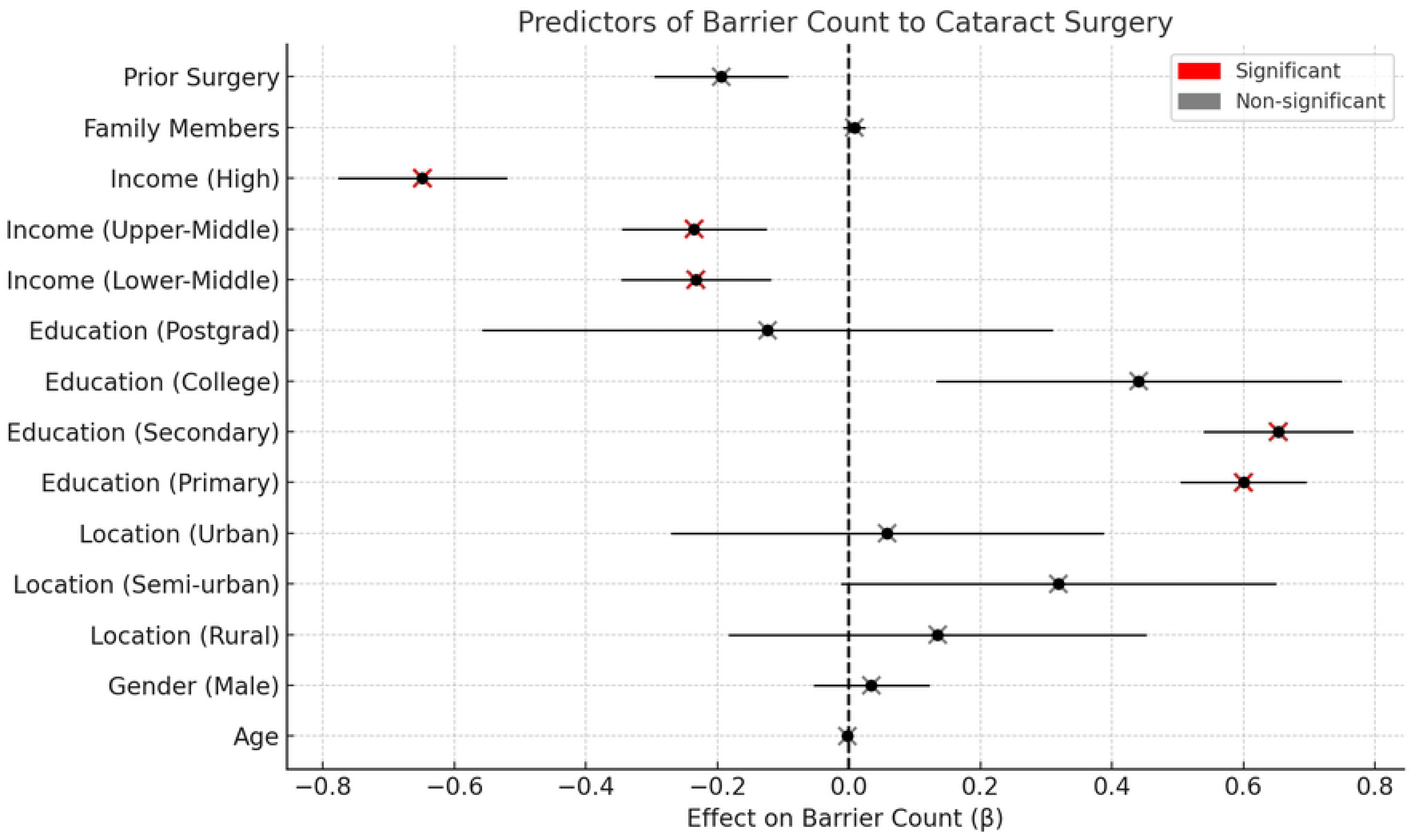
Predictors of Barrier Count to Cataract Surgery - Multivariable Regression Analysis. Forest plot displaying factors associated with the number of reported barriers to cataract surgery. Points represent adjusted effect estimates with confidence intervals shown as horizontal lines. Factors are ordered by their association strength, with positive values indicating more barriers and negative values indicating fewer barriers. Reference categories: Female gender, ethnic location, no formal education, and low-income group.

A multivariable regression analysis in table 4 reveals that patients with more reported barriers to cataract surgery had the strongest association with increased fear. Lower education levels, lower income, and female gender were also consistently associated with higher fear scores.

**Table 4:**
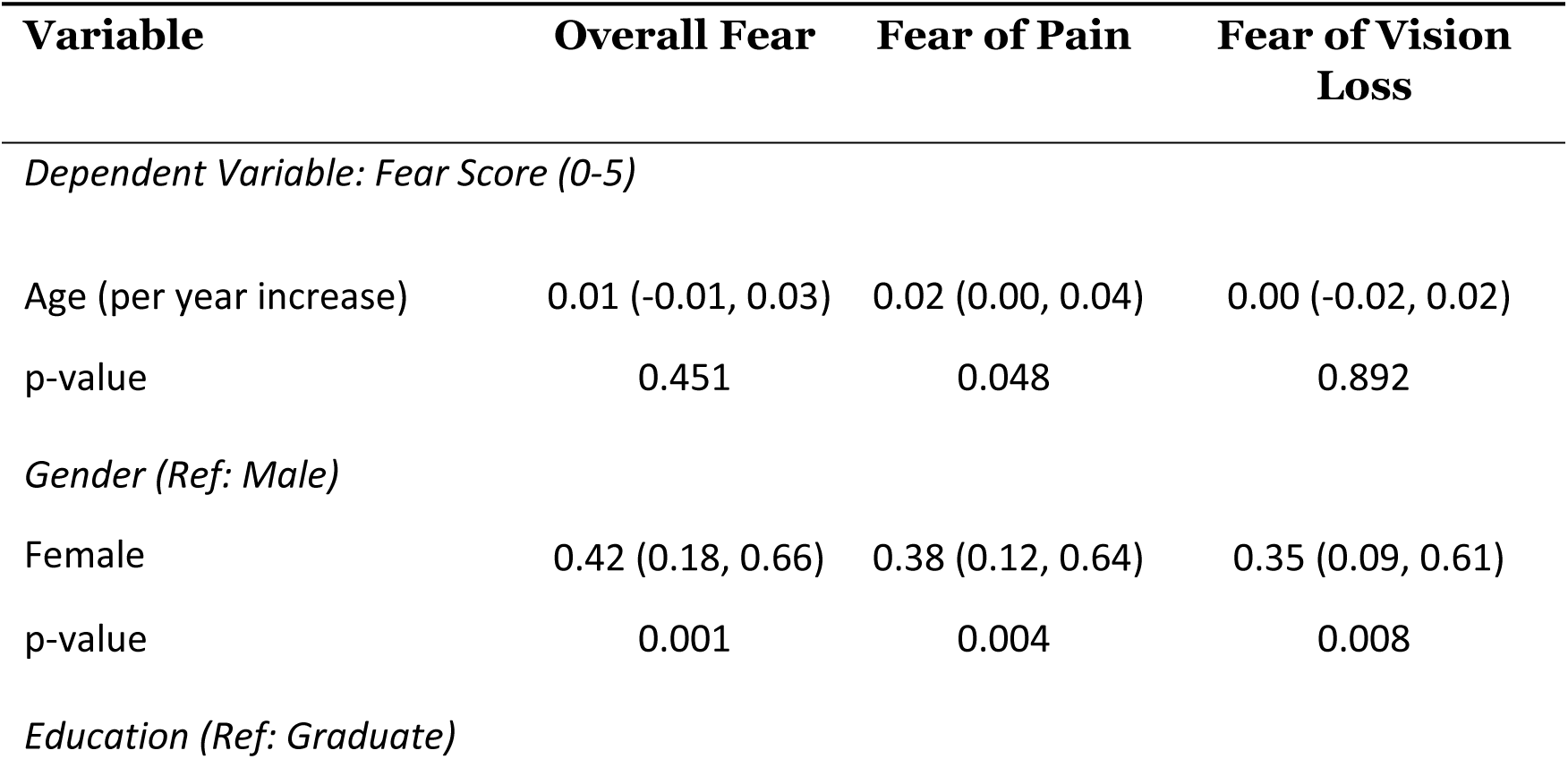
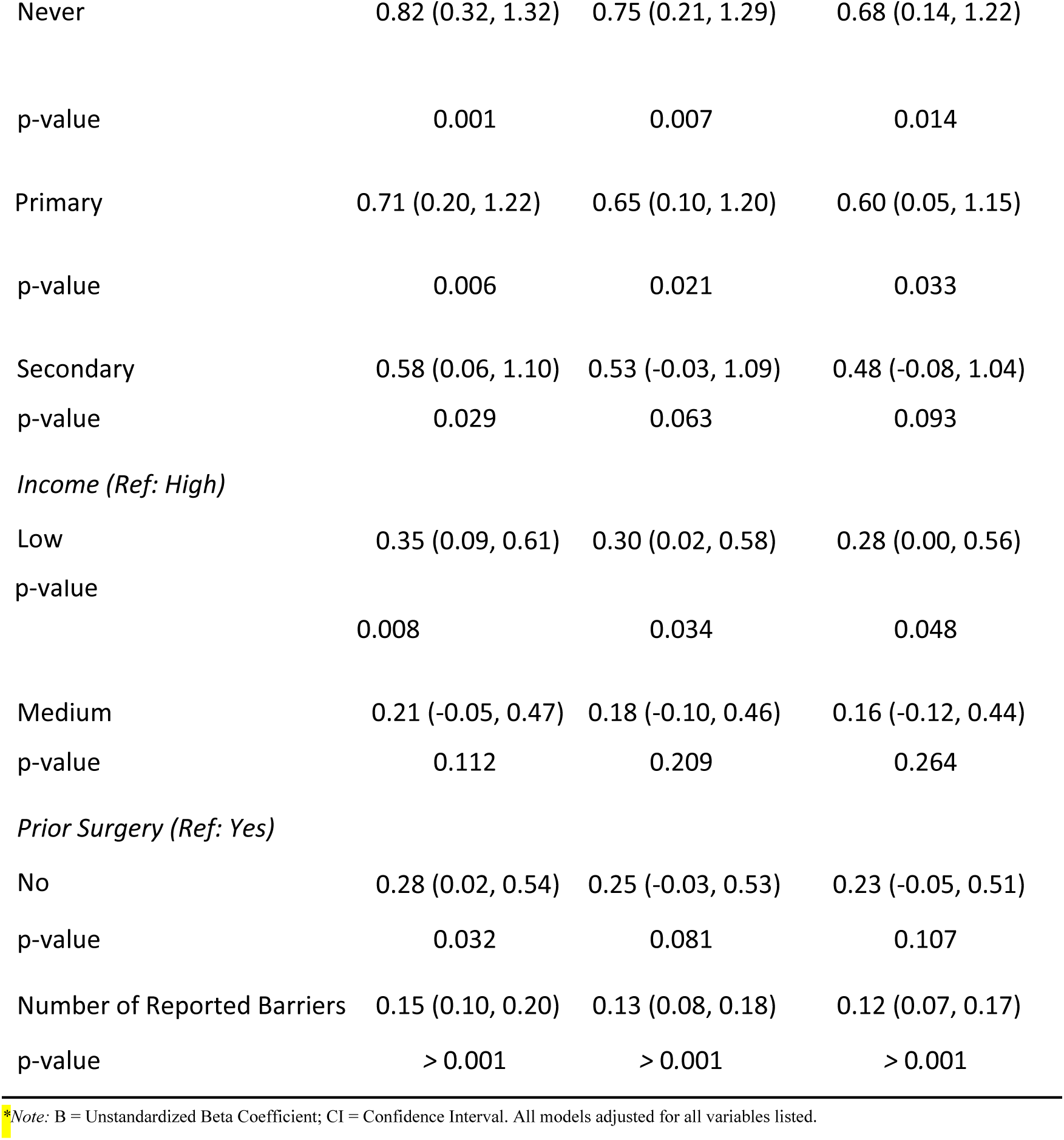
Multivariable Linear Regression Analysis of Factors Associated with Fear Scores (N=595)

| Variable | Overall Fear | Fear of Pain | Fear of Vision Loss |
| --- | --- | --- | --- |
| *Dependent Variable: Fear Score (0-5)* |  |  |  |
| Age (per year increase) | 0.01 (-0.01, 0.03) | 0.02 (0.00, 0.04) | 0.00 (-0.02, 0.02) |
| p-value | 0.451 | 0.048 | 0.892 |
| *Gender (Ref: Male)* |  |  |  |
| Female | 0.42 (0.18, 0.66) | 0.38 (0.12, 0.64) | 0.35 (0.09, 0.61) |
| p-value | 0.001 | 0.004 | 0.008 |
| *Education (Ref: Graduate)* |  |  |  |
| Never | 0.82 (0.32, 1.32) | 0.75 (0.21, 1.29) | 0.68 (0.14, 1.22) |
| p-value | 0.001 | 0.007 | 0.014 |
| Primary | 0.71 (0.20, 1.22) | 0.65 (0.10, 1.20) | 0.60 (0.05, 1.15) |
| p-value | 0.006 | 0.021 | 0.033 |
| Secondary | 0.58 (0.06, 1.10) | 0.53 (-0.03, 1.09) | 0.48 (-0.08, 1.04) |
| p-value | 0.029 | 0.063 | 0.093 |
| *Income (Ref: High)* |  |  |  |
| Low | 0.35 (0.09, 0.61) | 0.30 (0.02, 0.58) | 0.28 (0.00, 0.56) |
| p-value | 0.008 | 0.034 | 0.048 |
| Medium | 0.21 (-0.05, 0.47) | 0.18 (-0.10, 0.46) | 0.16 (-0.12, 0.44) |
| p-value | 0.112 | 0.209 | 0.264 |
| *Prior Surgery (Ref: Yes)* |  |  |  |
| No | 0.28 (0.02, 0.54) | 0.25 (-0.03, 0.53) | 0.23 (-0.05, 0.51) |
| p-value | 0.032 | 0.081 | 0.107 |
| Number of Reported Barriers | 0.15 (0.10, 0.20) | 0.13 (0.08, 0.18) | 0.12 (0.07, 0.17) |
| p-value | > 0.001 | > 0.001 | > 0.001 |

\*Note: B = Unstandardized Beta Coefficient; CI = Confidence Interval. All models adjusted for all variables listed.

Cataracts significantly disrupted the daily functioning of the majority of patients in this cohort. The most frequently reported impacts were on their ability to perform household tasks (e.g., cooking, cleaning; 47.9%), followed closely by challenges with mobility and safety (45.4%) and restrictions in leisure activities such as reading and watching television (44.5%). A substantial proportion of patients also reported adverse effects on their work or job performance (37.0%) and emotional well-being (32.8%), citing feelings of frustration, anxiety, and social isolation. Notably, only 10.1% of patients stated that their visual impairment had no significant impact.

## 4. Discussion

Visual impairment has a significant impact on health and quality of life, often contributing to dissatisfaction, disability, and reduced physical activity [19]. In the present study, the majority of patients reported difficulties with performing daily household chores (47.9%), physical movement (45.4%), and carrying out near-vision activities such as reading small print or viewing screens (44.5%). Some patients even indicated that their job performance and emotional well-being had deteriorated since the onset of a cataract.

When asked about barriers to cataract surgery, a clear majority of patients (72.3%) cited an inability to afford the procedure as their primary obstacle. This financial barrier was disproportionately shouldered by the most vulnerable segments of the population: those with the lowest levels of education, rural residents, and especially those with the lowest incomes. The cost of surgery has been identified as a significant barrier in multiple studies from low-income countries [10, 13, 20, 21]. Echoing our study findings, a population-based cross-sectional study in Papua New Guinea found that participants with inadequate education identified cost as a significant barrier while patients with secondary and tertiary education indicated time as the major constraint to cataract surgery [22]. Conversely, a study in Nepal indicated that even when participants were aware of the cataract treatment options, they could not access them due to financial constraints [13].

In our study, the majority of patients who identified work-related time constraints as a barrier were males and members of the younger population group, suggesting that concerns about loss of work opportunities contributed to delays in undergoing surgery. Patients often express difficulty affording cataract surgery, as lost wages and out-of-pocket expenses contribute to high medical costs [23, 24]. The financial burden of out-of-pocket expenses was greater among OPD patients in our study, as compared to camp patients, who received institutional support in the form of food, accommodation, and transportation. Patients with better socioeconomic conditions often delay surgery to secure a preferred surgeon or date, or to align the procedure with their travel or work schedules, thereby prolonging the waiting period [25].

### 4.1 Accessibility Barriers

Female and older participants (age>60) in the current study indicated “worry about surgical outcomes” and “a lack of escort” as barriers to seeking cataract surgery treatment on time. Similarly, the older population in Northeastern India cites the absence of an escort as a primary barrier to cataract surgery [30]. Besides, since male participants engaged in more income-generating activities, they had greater autonomy in decision-making than females. Additionally, women in traditional caregiver roles often lack sufficient time to access eye care services due to demanding household responsibilities and, in some contexts, a lack of permission or economic independence to travel for care outside their communities. Being confined to the home further restricts their access to health information and services [9, 26–28], which is consistent with our finding that men demonstrate greater knowledge of cataract treatments than women. The influence of ‘significant others,’ especially family and friends, plays a key role in shaping patients’ decisions [29]. Hence, women tend to rely more on men’s opinions while taking decisions concerning health. Comparably, a study in Nepal shows that men have greater access to cataract treatment, partly due to lower levels of awareness among women and the predominant role of male family members in making healthcare decisions [13]. In rural and underserved areas, lower educational attainment is often associated with poorer access to healthcare and a limited understanding of health information [13, 31].

The geographical barrier of transportation is predominant in our study. Literature suggests that patients from rural areas struggles to travel long distances to the clinic due to financial constraints. Elderly patients often rely on an escort to take them to the hospital for clinical procedures such as injections and pupil dilation [6].

Patients who delay seeing an eye doctor are more likely to postpone surgery and show up with advanced cataracts. In our study, such delays were frequently attributed to difficulties securing an appointment and the absence of available services, mirroring the findings of a cross-sectional survey conducted at a free eye clinic in Pennsylvania [32].

### 4.2 Socio-cultural Barriers

In the present study, patients with primary and secondary education reported the highest number of barriers. A study among the elderly population in Sri Lanka identified a lack of knowledge and awareness, socioeconomic factors, and misconceptions as the primary contributing factors to the low surgery rate, despite the high prevalence of cataracts [21]. Studies from rural China and systematic reviews also indicate that limited knowledge about cataracts and their treatment is a significant barrier to surgery uptake, with nearly one-third of patients declining surgery being unable to comprehend its necessity [33]. When patients lack understanding of IOL benefits, they often undervalue surgery, delay care until vision loss is severe, prioritize other needs, and rely on inaccurate non-medical information [34].

Patients with lower literacy levels are more likely to rely on their doctors for decision-making regarding IOL selection [34]. Physicians report that insufficient time to communicate the technology inside the IOLs hinder shared decision-making. Hence, better communication training and support from multidisciplinary nursing teams is required to educate patients and reduce physicians’ time burden [35].

Globally, fear of vision loss ranks first among all other fears of disabilities [36]. In the present study, patients with no prior surgery experience showed more fear compared to others. Since these patients are less familiar with ophthalmic procedures, they experience greater uncertainty about surgical outcomes [6]. Patients mostly fear that their vision might worsen with surgery, a fear often rooted in limited knowledge about cataracts, surgical procedures, and potential complications [10, 18, 21, 37]. Comparable to our findings, older patients in Nepal also reported fear of the procedure, worry regarding the outcome as the main barriers [38]. It is also evident from our findings that family support and behavioral changes play a crucial role in overcoming fear and accepting surgery.

The majority of our study participants expressed having a caregiver at home to assist them with medications. Qualitative studies also demonstrate that family support and having someone to accompany them are key factors motivating patients to undergo surgery [23, 39, 40]. In our study, more than half of the participants reported having a family member or friend with a positive surgical experience. Such positive accounts appear to foster reassurance, acting as a motivator for undergoing cataract surgery [26].

### 4.3 Limitations

This study exclusively included patients who underwent cataract surgery. The study was conducted in the hospital practice area; insightful findings could have been achieved if the study had been performed in slums or rural counterparts. A study in Africa found that asking patients about the reasons for not having surgery earlier can be extremely sensitive [41]. In such circumstances, patients are more likely to come up with an answer that does not embarrass them. There remains a possibility of social desirability bias in the current study, which can be mitigated through data triangulation and the systematic training of data collectors in the use of neutral language.

## 5. Conclusion

The study’s strength lies in its inclusion of a diverse population, thereby increasing its generalizability. The association between socio-demographics and specific barriers may help produce targeted interventions to improve access to cataract surgery. Public awareness can be raised through investing in intensive health promotion and partnering with local welfare organizations. Training medical staff to communicate the treatment options available for cataracts, and explaining the pros and cons of different available intraocular lenses to patients can also facilitate bridging the knowledge gap. More efforts should be made on patient counseling so that patients understand the nature, severity, and visual prognosis of their cataract.

## Data Availability

The data that support the findings of this study are available in Zenedo at https://doi.org/10.5281/zenodo.17336879

## Acknowledgment

The authors would like to express their sincere gratitude towards the patients and staff of Chattogram Lions Eye Institute and Hospital for their participation and support during data collection.

